# Serological response to COVID-19 vaccination in IBD patients receiving biologics

**DOI:** 10.1101/2021.03.17.21253848

**Authors:** Serre-Yu Wong, Rebekah Dixon, Vicky Martinez Pazos, ICARUS-IBD Working Group, Sacha Gnjatic, Jean-Frederic Colombel, Ken Cadwell

**Author notes:** **Corresponding author** Serre-Yu Wong, One Gustave L. Levy Place, Box 1069, New York, New York 10029. These authors share co-senior authorship (Jean-Frederic Colombel, Ken Cadwell). International study of COVID-19 Antibody Response Under Sustained immune suppression in IBD (ICARUS-IBD): Stephanie Gold, Drew Helmus, Jessica Anne Neil, Stela Sota, Kyung Ku Jang, Krystal Ching, Mericien Venzon, Xiaomin Yao, Lucie Bernard, Xin Chen, Reema Navalurkar, Michelle Mendiolaza, Pamela Reyes-Mercedes, Sara Nunez, Stephanie Stanley, Darwin Jimenez, Michael Tankelevich, Brianne Phillipe, Julio Ramos, Kevin Tuballes, Vanessa Barcessat, Natalia Herrera, Jack Satsangi, Kenji Watanabe, Séverine Vermeire, Flavio Steinwurz, Mark Silverberg, David T. Rubin, Giulia Roda, Walter Reinisch, Siew Chien Ng, James Lindsay, Jonas Halfvarson, Matthieu Allez, Vineet Ahuja, Maria Abreu.

## Abstract

**Objective:** The impact of medications on COVID-19 vaccine efficacy in IBD patients is unknown, as patients with immunosuppressed states and/or treated with immunosuppressants were excluded from vaccine trials. To address this, we evaluated serological responses to COVID-19 vaccination with the SARS-CoV-2 spike (S) mRNA BNT162b2 (Pfizer-BioNTech) and mRNA-1273 (NIH-Moderna) vaccines in IBD patients enrolled in an ongoing SARS-CoV-2 sero-survey at the Icahn School of Medicine at Mount Sinai in New York City.

**Design:** We obtained sera from 48 patients who had undergone vaccination with one or two vaccine doses. Sera were tested for SARS-CoV-2 anti-RBD total immunoglobulins and IgG (Siemens COV2T and sCOVG assays), anti-Spike IgG (in-house ELISA), and anti-nucleocapsid antibodies (Roche).

**Results:** All IBD patients (15/15) who completed two-dose vaccine schedules achieved seroconversion to high levels. Two IBD patients with history of COVID-19 infections and who were seropositive at baseline seroconverted to high levels after the first dose. Concurrent biologic use was 85% (41/48), including 33% of patients (16) on TNF antagonist monotherapy, 42% (17) on vedolizumab monotherapy, 6% (3) on vedolizumab combination therapy with thiopurine, and 8% (4) ustekinumab; 1 patient was receiving guselkumab for psoriasis. Three patients (6%) were on oral steroids at the time of vaccination.

**Conclusion:** IBD patients receiving biologics can seroconvert with robust serological responses after complete Pfizer-BioNTech and NIH-Moderna COVID-19 vaccination. In IBD-patients with previous SARS-CoV-2 seroconversion, a single dose of either vaccine can induce high index values, mirroring findings from the general population.

## Introduction

Inflammatory bowel disease (IBD) patients with Crohn’s disease (CD) and ulcerative colitis (UC), have been considered at increased risk of severe COVID-19 since they are often treated with immunosuppressive medications. Medications such as steroids and thiopurines particularly in combination therapy with TNF antagonists, though not TNF antagonist monotherapy, have been shown to contribute to the risk of severe COVID-19 in IBD patients^3,4^. Expert consensus advocates that IBD patients should be vaccinated against SARS-CoV-2^5^. Although data from other vaccine-preventable illnesses suggest attenuated responses in patients receiving TNF antagonists and immunomodulators^6^, the impact of medications on COVID-19 vaccine efficacy in IBD patients is unknown, as patients with immunosuppressed states and/or treated with immunosuppressants were excluded from vaccine trials. To address this, we evaluated serological responses to COVID-19 vaccination with the SARS-CoV-2 spike (S) mRNA BNT162b2 (Pfizer-BioNTech) and mRNA-1273 (NIH-Moderna) vaccines in IBD patients enrolled in an ongoing SARS-CoV-2 sero-survey at our institution^7^.

## Methods

Specimens were collected at routine infusion and outpatient clinic visits as part of an ongoing longitudinal SARS-CoV-2 sero-surveillance study of IBD patients at the Icahn School of Medicine at Mount Sinai. Patients were asked to report if they had at least one vaccination appointment between the first date of vaccine distribution in New York City on December 14, 2020, and February 12, 2021. Control cohorts included vaccinated (1) healthcare workers (HCW) without IBD who had completed both doses of either vaccine and (2) volunteers (PICR cohort) without IBD who underwent serial blood draws after vaccination. The study protocols are approved by the Icahn School of Medicine at Mount Sinai Institutional Review Board. IBD patient and HCW sera were analyzed using (1) the emergency use authorization (EUA) Siemens Healthineers COV2T and sCOVG assays which test for total immunoglobulins (Igs) and IgG, respectively, to the receptor binding domain (RBD) of the SARS-CoV-2 S protein and (2) the EUA Roche assay for antibodies to nucleocapsid protein. An in-house ELISA testing for IgG against full-length S protein was performed for IBD patients and both HCW and PICR controls. See Supplementary Methods for additional details.

## Results

Forty-eight IBD patients were included, including 23 CD and 25 UC patients (Supplementary Table 1). Most patients were receiving biologics at the time of vaccination (41 patients, 85%), including 16 (33%) TNF antagonist monotherapy, 17 (42%) vedolizumab monotherapy, 3 (6%) vedolizumab combination therapy with thiopurine, and 4 (8%) ustekinumab; 1 patient was receiving guselkumab for psoriasis. Three patients (6%) were on oral steroids at the time of vaccination. Five (10%) patients were on no medications. Controls, including 14 vaccinated HCWs (mean age 35.2, 50% female) and 29 vaccinated subjects in the PICR cohort (mean age 31.5, 39% female), were younger than the IBD cohort (p = 0.028 and p < 0.0001, respectively).

Participants received either Pfizer-BioNTech (IBD n = 23, HCW n = 11, PICR n = 20) or NIH-Moderna (IBD n = 25, HCW n = 3, PICR n = 9) vaccines. Thirty-three IBD patients had completed only the first dose, and 15 patients had completed both doses. All HCWs and 15 (52%) PICR controls completed both doses.

Three IBD patients (2 with previous COVID-19 and 1 with confirmed mild COVID-19 between dose 1 and 2) and one HCW (6% versus 7%) reported laboratory-confirmed COVID-19 infection by nasopharyngeal PCR or SARS-CoV-2 antibody testing after recovery. All other samples tested negative for anti-nucleocapsid antibodies. Pre-vaccine baseline samples obtained from 19 patients without a history of COVID-19 showed absence of SARS-CoV-2-specific antibodies by all assays, and 1 patient with prior COVID-19 had anti-RBD and anti-nuclecapsid antibodies at baseline. We were unable to collect baseline samples from the remaining patients due to enrollment after vaccination. Among 14 HCW controls, 1 (7%) had baseline sera drawn, and among PICR controls, 5 (17%) had baseline IgG reactivity to Spike protein due to prior infection.

All IBD patients (n = 15) who completed two-dose vaccine schedules seroconverted and achieved index levels which would have been high enough to qualify for convalescent plasma donation (ref) (Figure 1). Percent seroconversion by week is also shown. Two IBD patients with prior infection achieved high index values after a single vaccine dose, well above values achieved from natural SARS-CoV-2 infection (Figure 1a). Analysis of anti-S IgG levels of IBD patients compared to the PICR and HCW cohorts showed similar titers at most time points with a possible lag at 2-3 weeks post the first vaccine dose (p = 0.011) (Figure 1c).

**Figure 1.**
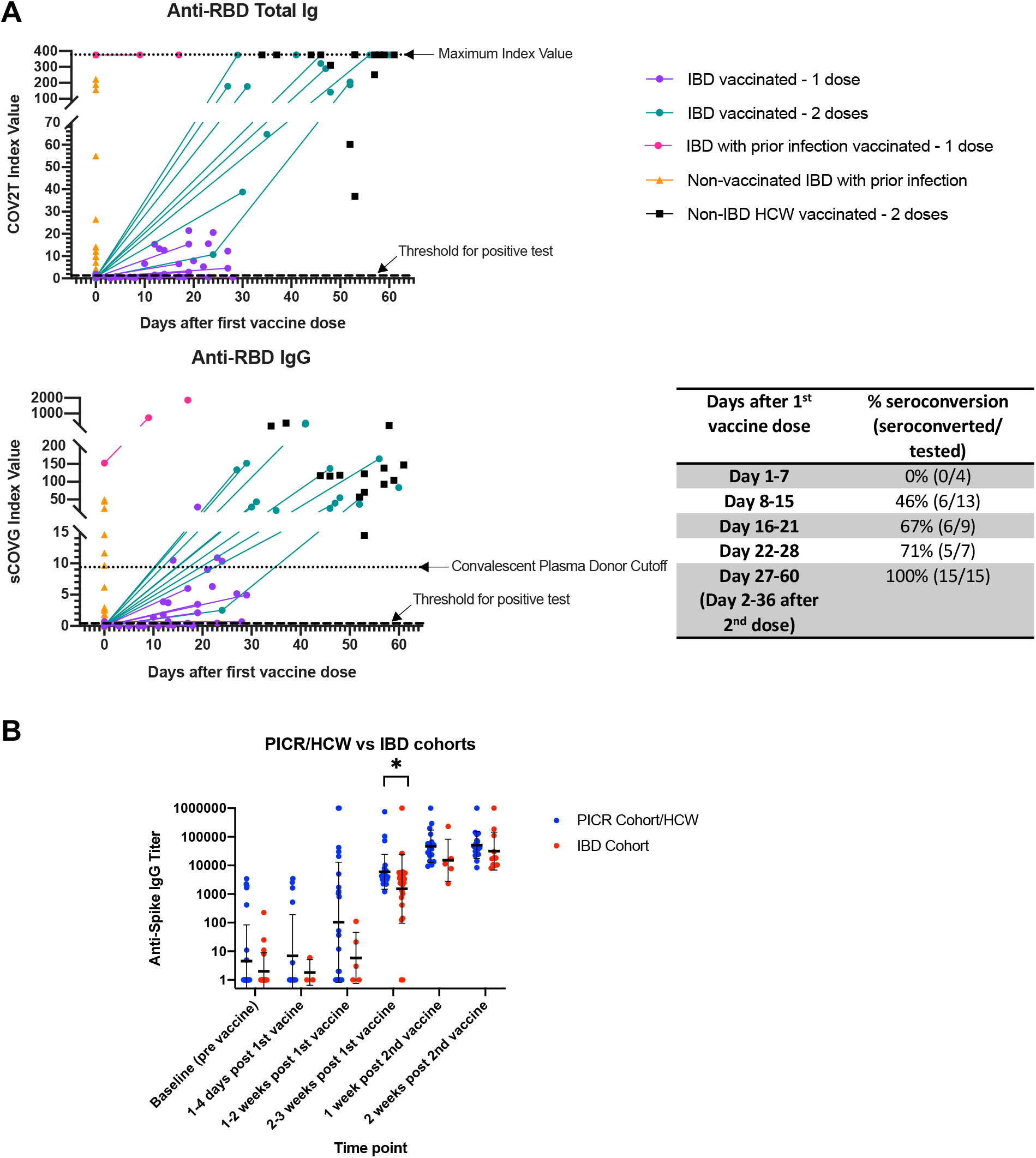
**Antibody response to SARS-CoV-2 immunization in IBD patients compared with healthcare worker controls. (A) Siemens COV2T and sCOVG testing for total Ig and IgG against SARS-CoV-2 RBD. Thresholds for positive tests and maximum index value for COV2T is shown by dotted lines as indicated. Percent seroconversion over time since fist vaccine dose in IBD patients is shown in the table. (C) Anti-Spike IgG results comparing IBD to PICR/HCW cohorts over time.**

Of the 15 patients who completed both COVID-19 vaccine doses, 5 were receiving TNF antagonist monotherapy, 9 vedolizumab monotherapy, and 2 no medications. Analyses of the effects of anti-TNF and vedolizumab monotherapy in these patients revealed no significant differences in index values for anti-RBD IgG. However, vedolizumab was associated with lower COV2T anti-RBD total Ig (0.02) and anti-S IgG (p = 0.0043) levels (Supplementary Figure 1).

## Discussion

We report a 100% seroconversion rate to complete Pfizer-BioNTech and NIH-Moderna COVID-19 vaccination in IBD patients on biologic monotherapy with robust serological responses. In IBD-patients with previous SARS-CoV-2 seroconversion, a single dose of either vaccine can induce high index values, mirroring findings from a recent HCW study^8^. We also found an association of lower antibody levels in patients with vedolizumab. This finding is of unclear clinical significance and warrants further investigation, as this result could have been affected by timing, vaccine, or clinical characteristics such as age.

This is the first data of serological responses to COVID-19 vaccines in IBD patients with detailed analysis of antibodies to both nucleocapsid and RBD/spike proteins. The study is limited to our single center experience. In addition, there were differences in time to blood collections between IBD patients and controls. Despite these limitations, our results support the consensus recommendation for IBD patients to receive COVID-19 vaccines when available^5^. Our study also highlights the need for future studies to assess earlier time points and longitudinal studies to assess full immune responses, including cell-mediated responses, effects of other medications, and clinical efficacy of different vaccine types to guide optimal dosing regimens and/or vaccine choice in IBD patients.

## Data Availability

Data will be made available upon request.

## Conflict of Interest

Sacha Gnjatic: Research grants from Bristol-Myers Squibb, Genentech, Immune Design, Agenus, Janssen R&D, Pfizer, Takeda, and Regeneron; advisory roles for Merck, Neon Therapeutics and OncoMed. Jean-Frederic Colombel: Research grants from AbbVie, Janssen Pharmaceuticals and Takeda; receiving payment for lectures from AbbVie, Amgen, Allergan, Bristol-Myers Squibb Company, Ferring Pharmaceuticals, Shire, Takeda and Tillots; has received consulting fees from AbbVie, Amgen, Arena Pharmaceuticals, Boehringer Ingelheim, Bristol-Myers Squibb Company, Celgene Corporation, Celltrion, Eli Lilly, Enterome, Ferring Pharmaceuticals, Genentech, Gilead, Iterative Scopes, Ipsen, Immunic, lmtbio, Inotrem, Janssen Pharmaceuticals, Landos, LimmaTech Biologics AG, Medimmune, Merck, Novartis, O Mass, Otsuka, Pfizer, Shire, Takeda, Tigenix, Viela bio; and holding stock options in Intestinal Biotech Development. Ken Cadwell: Research funding from Pfizer, Takeda, and Abbvie; consulted for or received an honorarium from Puretech Health, Genentech, and Abbvie; and holds U.S. patent 10,722,600 and provisional patent 62/935,035. The remaining authors disclose no conflicts.

## Funding

This work was supported by the Helmsley Charitable Trust and Cure for IBD. This effort was supported by the Serological Sciences Network (SeroNet) of the National Cancer Institute, National Institutes of Health, under Contract No. 75N91019D00024, Task Order No. 75N91020F00003. The content of this publication does not necessarily reflect the views or policies of the Department of Health and Human Services, nor does mention of trade names, commercial products or organizations imply endorsement by the U.S. Government. Sacha Gnjatic was supported by NIH grants CA224319 and DK124165. This work was supported in part through the computational resources and staff expertise provided by Scientific Computing at the Icahn School of Medicine at Mount Sinai. Supported by grant UL1TR001433 from the National Center for Advancing Translational Sciences, National Institutes of Health. The funders had no role in the design and conduct of the study; collection, management, analysis, and interpretation of the data; preparation, review, or approval of the manuscript; and decision to submit the manuscript for publication.

## Authorship Contributions

Serre-Yu Wong (Conceptualization: Lead; Data curation: Equal; Formal analysis: Equal; Funding acquisition: Lead; Investigation: Equal; Methodology: Equal; Project administration: Equal; Supervision: Lead; Visualization: Lead; Writing – original draft: Lead; Writing – review & editing: Equal). Rebekah Dixon (Conceptualization: Equal; Data curation: Equal; Investigation: Equal; Methodology: Equal; Project administration: Equal; Writing – review & editing: Equal). Vicky Martinez Pazos (Investigation: Equal; Methodology: Equal; Lead; Writing – review & editing: Supporting). Sacha Gnatic (Conceptualization: equal; Data curation: Equal; Formal analysis: Equal; Methodology: Equal; Visualization: Equal; Writing – review & editing: Equal). Jean-Frederic Colombel (Conceptualization: Equal; Formal analysis: Equal; Funding acquisition: Equal; Methodology: Supporting; Writing – review & editing: Equal). Ken Cadwell (Conceptualization: Equal; Formal analysis: Equal; Funding acquisition: Equal; Methodology: Equal; Writing – review & editing: Equal).

## Acknowledgements

The authors wish to thank The Mount Sinai Therapeutic Infusion Center team and Yamilka Costanza for their contributions to patient recruitment; James Freeman, Justin Conklin, Neil Birmingham, Don Chalfin, Ross Molinaro, Kim Wilson, and Siemens for their support for serological testing; Gustavo-Martinez-Delgado and Louis Cohen for laboratory support; Miriam Merad for valuable input on the study design.

Author names in bold designate shared co-first authorship.

## Notes

### Author Declarations

The Institutional Review Board of the Mount Sinai School of Medicine reviewed and approved the SARS-CoV-2 exposure in patients with altered immune function study (IRB 20-03464).

