## Supplementary Table, Figure, and Methods for "Serological response to COVID-19 vaccination in IBD patients receiving biologics"

### Supplemental Table 1. Baseline characteristics of vaccinated individuals

| Characteristic | | Vaccinated IBD Patients n= 48 | Vaccinated Non- IBD HCW[a] (Controls) n= 14 | Vaccinated PICR Cohort (Controls) n = 29 | p-value |
|---|---|---|---|---|---|
| Age, mean, years | | 48.8 | 35.2 | | 0.028 |
| | | | | 31.5 | <0.0001 |
| Gender, female, n (%) | | 24 (50%) | 7 (50%) | | 1.000 |
| | | | | 11 (39%) | 0.43 |
| Race, n(%) | White | 42 (88%) | 10 (71%) | | 0.21 |
| | Non-White | 6 (12%) | 4 (29%) | | |
| | | | | 18 (62%) | 0.02 |
| | | | | 11 (38%) | |
| Type of IBD, n (%) | Crohn’s Disease | 23 (48%) | - | - | - |
| | Ulcerative Colitis | 25 (52%) | - | - | - |
| IBD medications, n (%) | Infliximab mono | 14 (29%) | - | - | - |
| | Adalimumab mono | 2 (4%) | - | - | - |
| | Vedolizumab mono | 17 (42%) | - | - | - |
| | Vedolizumab + Immunomodulator | 3 (6%) | | | |
| | Ustekinumab | 4 (8%) | - | - | - |
| | Tofacitinib | 1 (2%) | - | - | - |
| | Steroids, Oral | 3 (6%) | - | - | - |
| | Immunomodulator | 3 (6%) | - | - | - |
| | Mesalamine | 11 (23%) | - | - | - |
| | Biologic, any[b] | 41 (85%) | - | - | - |
| | No IBD medications | 5 (10%) | 14 (100%) | - | - |
| Known prior COVID-19 infection, n (%) | | 3 (6%) | 1 (7%) | - | 1.000 |
| Vaccine type | Pfizer | 23 (48%) | 11 (79%) | | |
| | Moderna | 25 (52%) | 3 (21%) | | 0.066 |
| | | | | 20 (69%) | |
| | | | | 9 (31%) | 0.12 |
| Time to blood collection after $1^{st}$ dose, median (range) | | 14 (3-28) | - | 9 (1-40) | <0.0001 |
| Time to blood collection after $2^{nd}$ dose, median (range) | | 18 (2-36) | 30 (7-37) | | 0.0001 |
| | | | | 8 (6-18) | <0.0001 |
| Vaccine reaction, yes, n/total respondents (%) | | 29/36 (81%) | 13/14 (93%) | - | 0.024 |
| | Severe reaction | 0 (0%) | 0 (0%) | - | |
| | Local arm pain/swelling/rash | 19 (53%) | 9 (64%) | - | 0.68 |
| | Myalgia | 12 (33%) | 8 (57%) | - | 0.22 |
| | Arthralgia | 1 (3%) | 3 (21%) | - | 0.11 |
| | Fatigue | 14 (39%) | 7 (50%) | - | 0.69 |
| | Headache | 9 (25%) | 6 (43%) | - | 0.37 |
| | Fever/subjective fever | 12 (33%) | 2 (14%) | - | 0.32 |
| | Chills | 8 (22%) | 2 (14%) | - | 0.81 |
| | GI symptoms[c] | 4 (11%) | 0 (0%) | - | 0.47 |
| | Other rash[d] | 1 (3%) | 0 (0%) | - | 0.53 |
| | Other localized pain[e] | 3 (6%) | 0 (0%) | - | 0.65 |

[a] Healthcare worker (HCW)
[b] One patient received guselkumab (anti-IL23) for psoriasis but takes no medications for UC.
[c] GI symptoms included nausea, diarrhea, and self-described Crohn’s flare.
[d] Stomach rash
[e] Pain localized to breast, knee, and leg

**Supplementary Methods**

*Patients.* Patients enrolled in the study reported COVID-19 vaccination status and reported dates and types of vaccination, vaccine reactions by self-reporting, current medications, COVID-19 testing and illness history by an online survey, email responses, and follow-up phone calls. Age, sex, race, type of IBD, and medications were confirmed by medical record.

*Sample processing*. Blood specimens were collected in SST tubes, allowed to clot and centrifuged at 1100-1300g for 20 minutes at room temperature. The specimens were aliquoted into sterile cryovials and stored immediately at -80˚C until testing.

*Serology testing.* The Siemens COV2T chemiluminescence-based assay measures total antibodies to the SARS-CoV-2 RBD of the S protein, and the sCOVG is a semi-quantitative assay for anti-RBD immunoglobulin (Ig)G [refs]. While both the COV2T and sCOVG are semi-quantitative assays, at the time of this writing only the sCOVG assay has EUA for semi-quantitative index value results. An index value of 1 equals a positive test. These tests are expected to detect seroconversion both to SARS-CoV-2 infection and vaccines which are designed to deliver Spike protein antigen. In order to distinguish serological response secondary to vaccine versus natural infection, all sera were additionally tested by the EUA Roche assay for antibodies (IgG) to nucleocapsid protein, which is not targeted by currently-approved vaccines in the U.S and would therefore be expected only to be positive in sera from individuals with SARS-CoV-2 infection. For the in-house ELISA to Spike S protein, sera were serially diluted from 1/100 to 1/6400 and results were expressed as reciprocal titers based on the predicted dilution at which a linear extrapolation of the titration curve meets a cutoff determined from a healthy donor serum pool. A titer ≥100 was considered positive, and 4x titer increase from baseline as significant.

*Statistical methods.* Statistical analysis was performed using R v3.5.3 and GraphPad Prism v9. For categorical covariates, p-values were calculated using the chi-square test with Yates continuity correction. For continuous covariates, p-values were calculated using Student's t-test, Mann-Whitney test, or Wilcoxon test.

**Supplementary Figure 1. Comparison of SARS-CoV2 antibodies in IBD patients who received two COVID-19 vaccine doses by medication.**

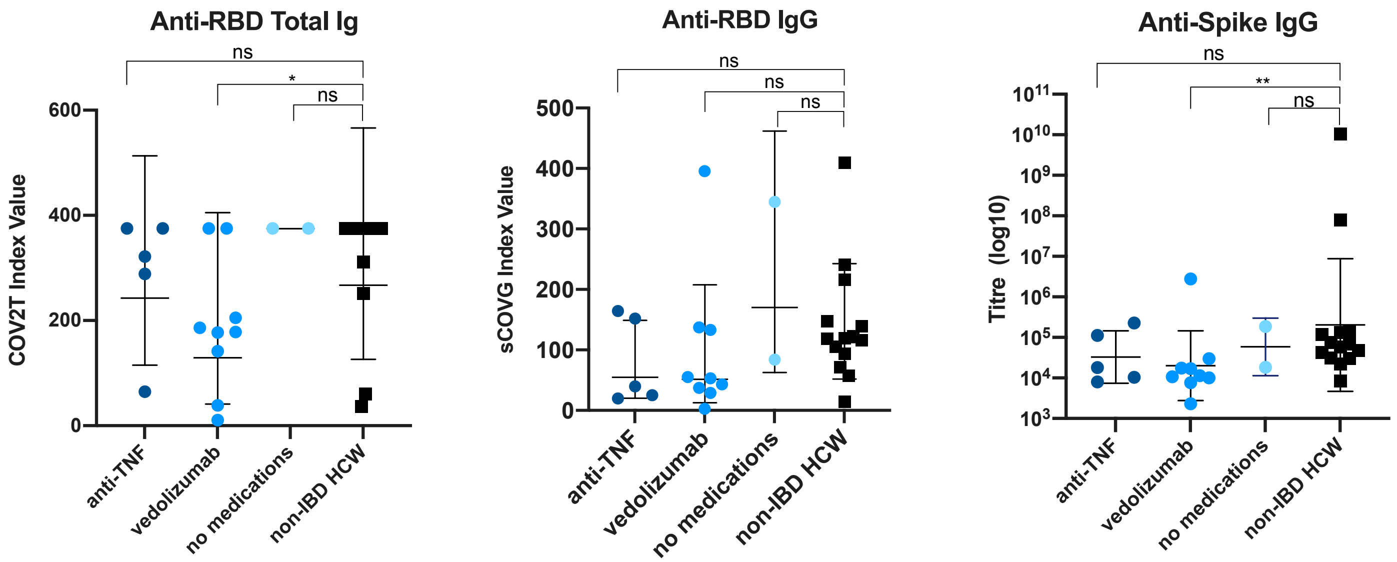
